# Influenza Vaccination and Major Cardiovascular Risk: A Systematic Review and Meta-Analysis of Clinical Trials Studies

**DOI:** 10.1101/2023.08.04.23293585

**Authors:** Fatemeh Omidi, Moein Zangiabadian, Amir Hasehm Shahidi Bonjar, Mohammad Javad Nasiri, Tala Sarmastzadeh

**Affiliations:** Department of Cardiology, Imam Hossein Hospital, Shahid Beheshti University of Medical Sciences, Tehran, Iran; School of Medicine, Shahid Beheshti University of Medical Sciences, Tehran, Iran

**Keywords:** Influenza vaccination, cardiovascular events, myocardial infarction, cardiovascular death, stroke, risk reduction

## Abstract

**Background:** Cardiovascular events remain a substantial global health concern, necessitating innovative strategies for prevention. This study aims to assess the potential impact of influenza vaccination on major cardiovascular events.

**Methods:** A search of the medical English literature was conducted using PubMed/MEDLINE, EMBASE, and the Cochrane CENTRAL up to 1 August 2023. Meta-analysis and stratified analyses were performed to investigate specific outcomes, including myocardial infarction (MI), cardiovascular death, and stroke. Pooled relative risks (RR) along with their 95% confidence intervals (CI) were calculated to evaluate the associations.

**Results:** A comprehensive analysis was conducted on a total of 9059 patients, with 4529 patients receiving the influenza vaccine and 4530 patients receiving a placebo. Among patients who received the influenza vaccine, a notable reduction in the occurrence of major cardiovascular events was observed, with 517 cases compared to 621 cases in the placebo group (RR, 0.70; 95%CI, 0.55-0.91). The stratified analysis revealed a decreased risk of MI in vaccinated patients (RR, 0.74; 95%CI, 0.56-0.97) and a significant reduction in cardiovascular death events (RR, 0.67; 95% CI 0.45–0.98).

**Conclusion:** This study provides compelling evidence that influenza vaccination is associated with a decreased risk of major cardiovascular events, particularly myocardial infarction, and cardiovascular death. These findings highlight the potential of influenza vaccination as an adjunctive strategy in cardiovascular disease prevention. Further research and exploration of underlying mechanisms are warranted to elucidate the observed beneficial effects.

## Introduction

The significance of influenza vaccination has long been acknowledged in preventing seasonal flu infections, particularly in high-risk populations [1-3]. There are strong recommendations for influenza vaccination where six months and older people especially adults should be vaccinated against influenza [3].

Cardiovascular diseases (CVDs) including hypertensive heart disease, ischemic heart disease, atrial fibrillation cerebrovascular disease like stroke, endocarditis, peripheral vascular disease, and other related cardiovascular diseases are a leading cause of morbidity and mortality worldwide [4].

Differing viewpoints exist regarding the impact of influenza vaccination on CVDs. While certain observational investigations suggest a favorable correlation between influenza vaccination and the reduction in occurrences of cardiovascular incidents like acute myocardial infarction (MI) [5, 6], contrasting epidemiological studies propose the limited efficacy of influenza vaccines [6].

Consequently, a comprehensive and updated study becomes imperative. This updated meta-analysis seeks to assess whether a connection exists between influenza vaccination and a decreased likelihood of experiencing cardiovascular events.

## Methods

### Search Strategy

A search of the medical English literature was conducted using PubMed/MEDLINE, EMBASE, and the Cochrane CENTRAL up to 1 August 2023. A thorough review was conducted on randomized controlled trials (RCTs) exploring the potential link between influenza vaccination and the subsequent risk of developing CVDs.

The search terms were: myocardial disease and influenza vaccines (Table S1). This study adhered to the Preferred Reporting Items for Systematic Reviews and Meta-Analyses statement (PRISMA) for its design and reporting (Prospero pending ID: 450694) [4].

### Study Selection

The records obtained from the database searches were amalgamated, and redundancy was addressed through employment of EndNote X7 (Thomson Reuters, Toronto, ON, Canada). Subsequently, two evaluators (MZ and MJN) carried out individual assessments of the records, considering both their title/abstract and full text, in order to exclude any entries that did not align with the study’s objectives.

The studies included in the analysis met the following criteria:

*Participants:* The patients with a diagnosis of CVDs.

*Intervention:* Patients with CVDs who received influenza vaccine.

*Comparison:* Patients with CVDs who received a placebo.

*Outcome:* Lower risk of cardiovascular events.

Conference abstracts, case reports, and studies comparing high and low doses of influenza vaccination were excluded.

### Data Extraction

Data extraction was conducted by two investigators (MZ and MJN) and recorded in a Microsoft Excel spreadsheet (XP professional edition; Microsoft Corp, Redmond, WA). The extracted information encompassed various key elements, including the first author’s name, publication date, study type, definitions of cases and controls, the number of cases and controls, and the outcomes of the studies.

### Quality Assessment

The quality assessment of the included studies was conducted by two reviewers (MJN and MZ) using the Cochrane tool [5]. In case of any discrepancies, a third reviewer (FO) was involved. This instrument encompasses a range of domains, which encompass random sequence generation, allocation concealment, blinding of both participants and personnel, blinding of assessors for outcomes, completeness of outcome data, and additional considerations such as selective reporting and potential sources of bias. Each study was categorized as having a low risk of bias when no concerns regarding bias were identified, a high risk of bias when there were concerns about bias or an unclear risk of bias when there was insufficient information available.

### Statistical Analysis

The combined outcomes were presented as risk ratios along with corresponding 95% confidence intervals (CIs). To evaluate the level of variability among the studies, both the I2 value and p-value were employed. In cases where minimal statistical heterogeneity was observed (I2 ≤ 50% or p ≥ 0.1), the fixed-effect model was applied. Conversely, if substantial inter-study heterogeneity was identified (I2 > 50% or p < 0.1), the random-effects model was utilized. Assessment of between-study heterogeneity involved the application of Cochran’s Q test and the I2 statistic. To assess potential publication bias, Begg’s test was employed, where a significance level of p<0.05 indicated noteworthy publication bias. All analytical procedures were conducted using Comprehensive Meta-Analysis Software, Version 3.0 (Biostat, Englewood, NJ).

## Results

Out of the 275 articles initially identified, five studies comprising 9,059 patients met the predetermined inclusion criteria, as illustrated in Figure 1 [6-10]. The key characteristics of these selected studies are summarized in Table 1. Among the participants, a total of 4,529 individuals were randomly assigned to receive a standard intramuscular influenza vaccination, while 4,530 were assigned to receive an intramuscular placebo. The average age of the participants was 61.3 years, and they were followed up for an average duration of 9 months. The majority of the trials exhibited robust practices in terms of randomization, allocation concealment, and masking, aligning with the Cochrane criteria for high quality (low risk of bias), except for one trial (Table 2).

**Figure1.**
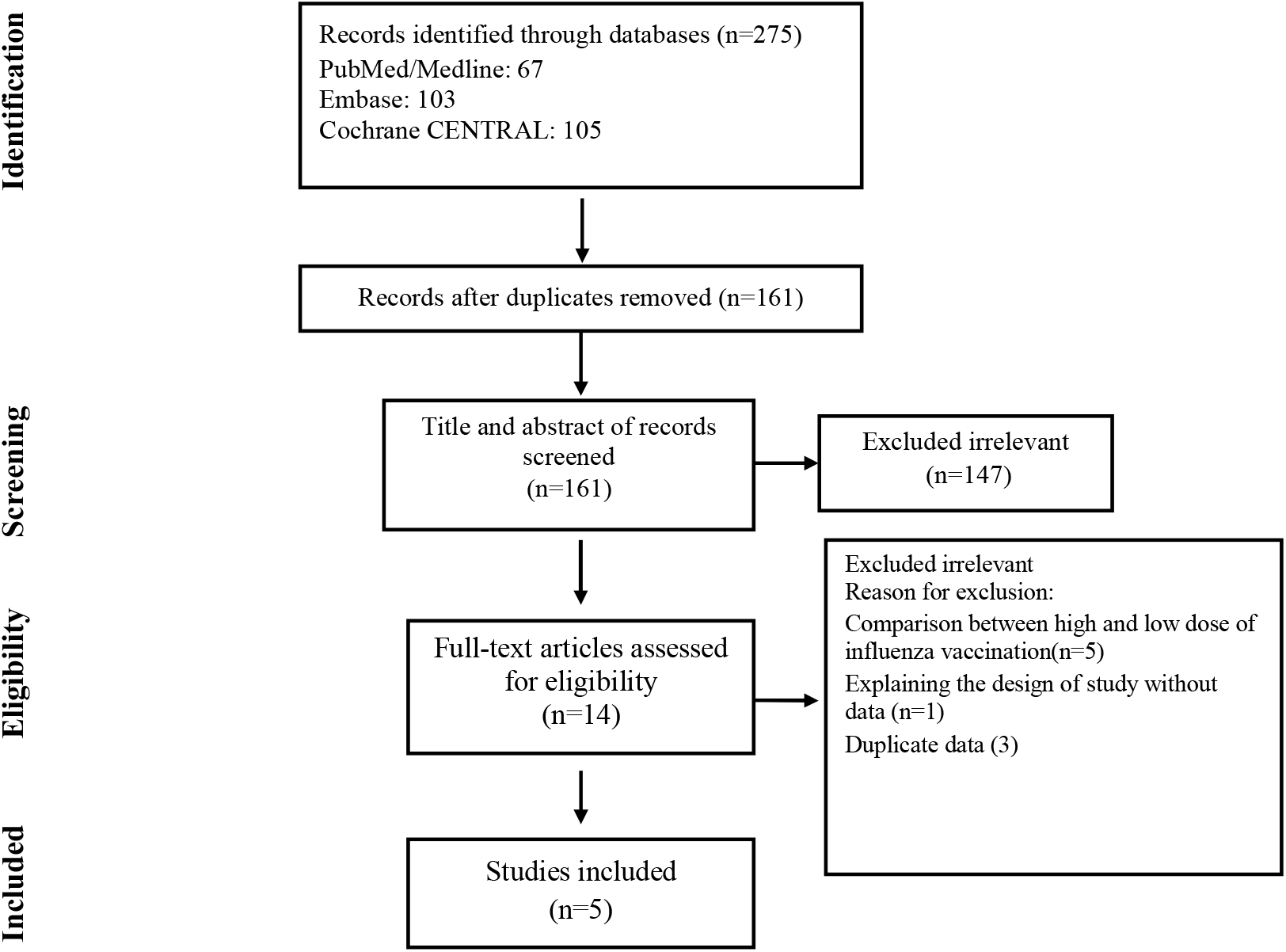
Flow chart of study selection for inclusion in the systematic review and meta-analysis.

**Table1.** Characteristic of Included Studies.

| **First author** | **Published year** | **Country** | **Mean Age** | **Definition of case (Number of participants)** | **Definition of control (Number of participants)** | **Outcome** |
| --- | --- | --- | --- | --- | --- | --- |
| Loeb [6] | 2022 | Multi-center | 57 | Vaccinated patients with heart failure and NYHA functional class II, III, and IV (2560) | Unvaccinated patients with heart failure and NYHA functional class II, III, and IV (2569) | IHD, Stroke, Cardiovascular death |
| Frobert [7] | 2021 | Multi-center | 60 | Vaccinated patients with MI (1272) | Unvaccinated Participants with MI (1260) | IHD, Stroke, Cardiovascular death |
| Phrommintikul [8] | 2011 | Thailand | 66 | Vaccinated patients with the acute coronary syndrome (n = 221) | Unvaccinated patients with the acute coronary syndrome (n = 218) | IHD, Stroke, Cardiovascular death |
| Ciszewski [9] | 2008 | Poland | 60 | Vaccinated patients with coronary artery disease (n = 325) | Unvaccinated patients with coronary artery disease (n = 333) | IHD, Cardiovascular death |
| Gurfinkel [10] | 2002 | Argentina | 65 | Vaccinated patients with MI or planned stenting (n = 151) | Unvaccinated patients with MI or planned stenting (n = 150) | IHD, Cardiovascular death |

**Table 2.** Quality Assessment.

| Author | Random sequence generation | Allocation concealment | Blinding of participants and personnel | Blinding of outcome assessment | Incomplete outcome data |
| --- | --- | --- | --- | --- | --- |
| Loeb | Low | Low | Low | Low | Low |
| Frobert | Low | Low | Low | Low | Low |
| Phrommintikul | Low | Low | Unclear | Unclear | Low |
| Ciszewski | Low | Low | Low | Unclear | Low |
| Gurfinkel | Low | High | Unclear | Unclear | Low |

### Pooled Major Cardiovascular Events

Within the cohort of 4,529 patients who underwent influenza vaccination, a total of 517 individuals experienced significant cardiovascular events, in contrast to 621 cases among the 4,530 patients who were administered a placebo. The calculated risk ratio (RR) was 0.70, indicating a noteworthy reduction in risk, accompanied by a 95% confidence interval (CI) of 0.55 to 0.91 (as depicted in Figure 2).

**Figure2.**
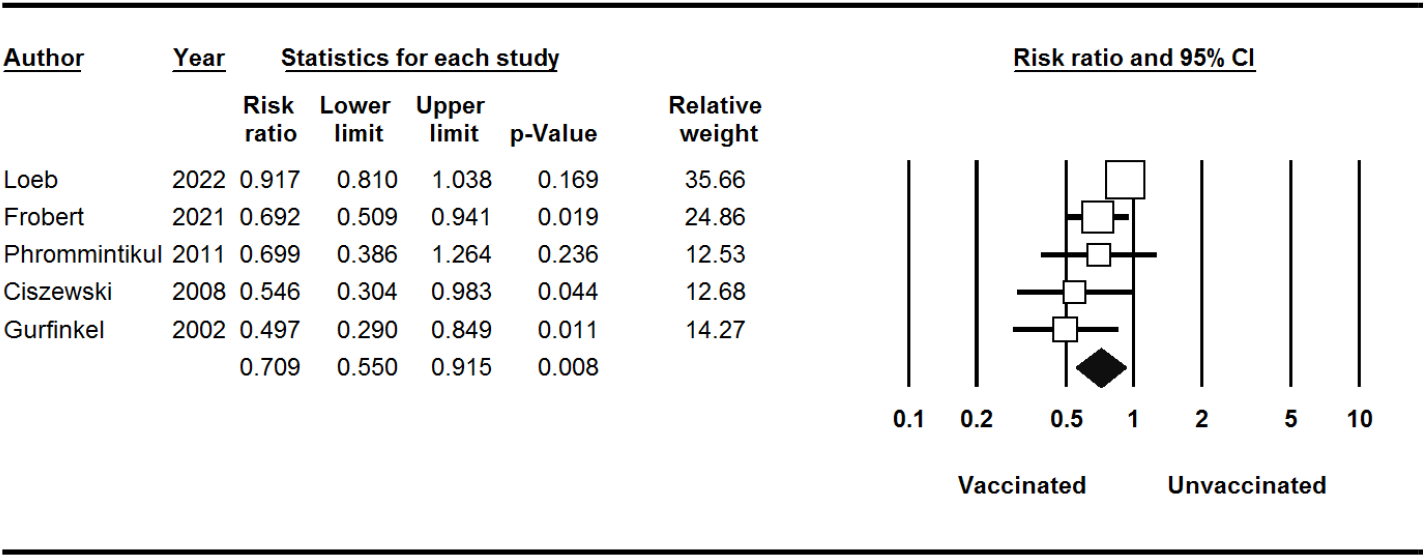
Major Cardiovascular Events for Influenza Vaccine Vs Control.

### Pooled Stratified Analysis

Stratified analyses are shown in Table 3. MI occurred in 86 of the 4529 patients who received influenza vaccine compared with 116 of the 4530 patients who received placebo (RR, 0.74; 95%CI, 0.56-0.97) (Figure 3). The RR of developing cardiovascular death events after influenza vaccination was 0.67 (95% CI 0.45– 0.98) (Figure 4). Thus, influenza vaccination significantly decreased the risk of cardiovascular death (P-value = 0.04). there was not any significant association between influenza vaccination and risk of stroke (P-value = 0.7) (Figure 5).

**Table 3.** Stratified Pooled Analysis.

| Outcome | No. of study | No. of patients | Risk ratio<br>(95 % CI)/(p-value) | Heterogeneity<br>$I^2$ (%) / p-value | Begg test<br>p-value |
| --- | --- | --- | --- | --- | --- |
| Major Cardiovascular Events | 5 | 9059 | 0.70 (0.55-0.91) / 0.00 | 58.16 / 0.04 | 0.80 |
| **Stratified Analysis** |  |  |  |  |  |
| Myocardial Infarction | 5 | 9059 | 0.74 (0.56-0.97) / 0.03 | 0.00 / 0.71 | 1.00 |
| Stroke | 3 | 8100 | 1.05 (0.72-1.54) / 0.77 | 0.00 / 0.65 | 1.00 |
| Cardiovascular Death | 5 | 9059 | 0.67 (0.45-0.98) / 0.04 | 51.19 / 0.08 | 0.80 |

**Figure3.**
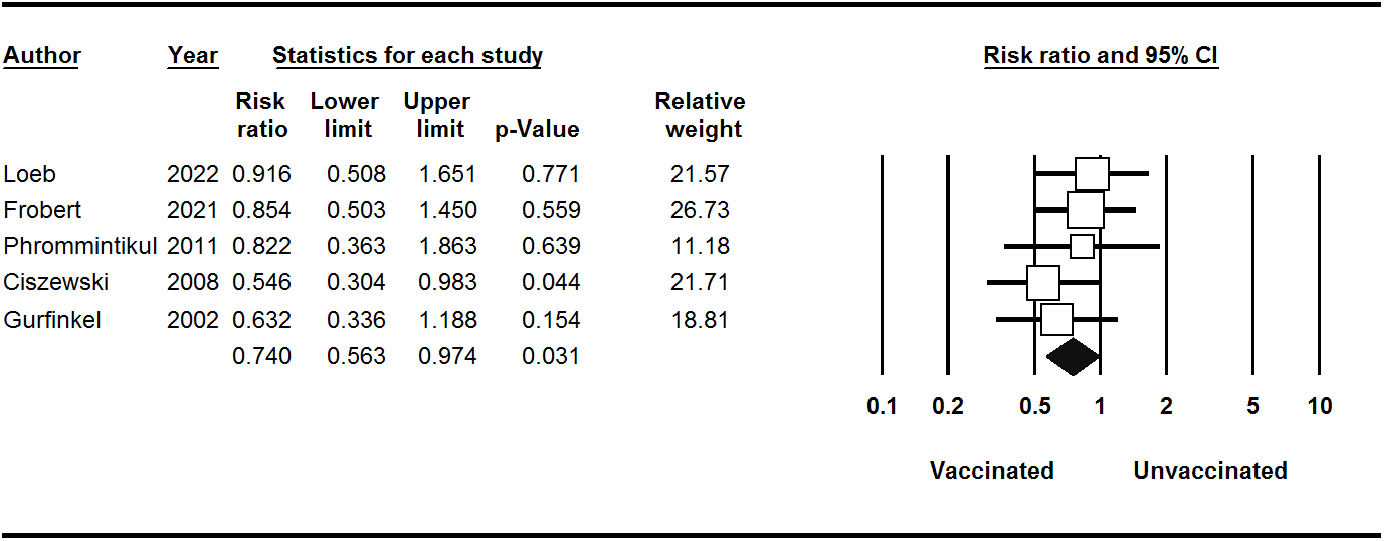
MI Events for Influenza Vaccine Vs Control.

**Figure4.**
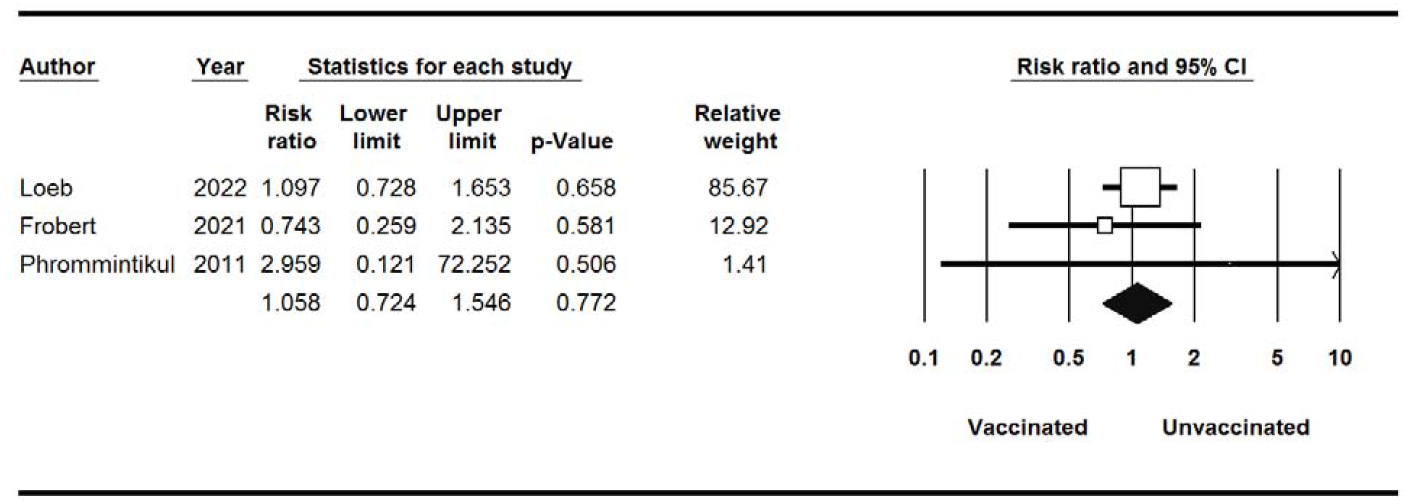
Stroke Events for Influenza Vaccine Vs Control.

**Figure5.**
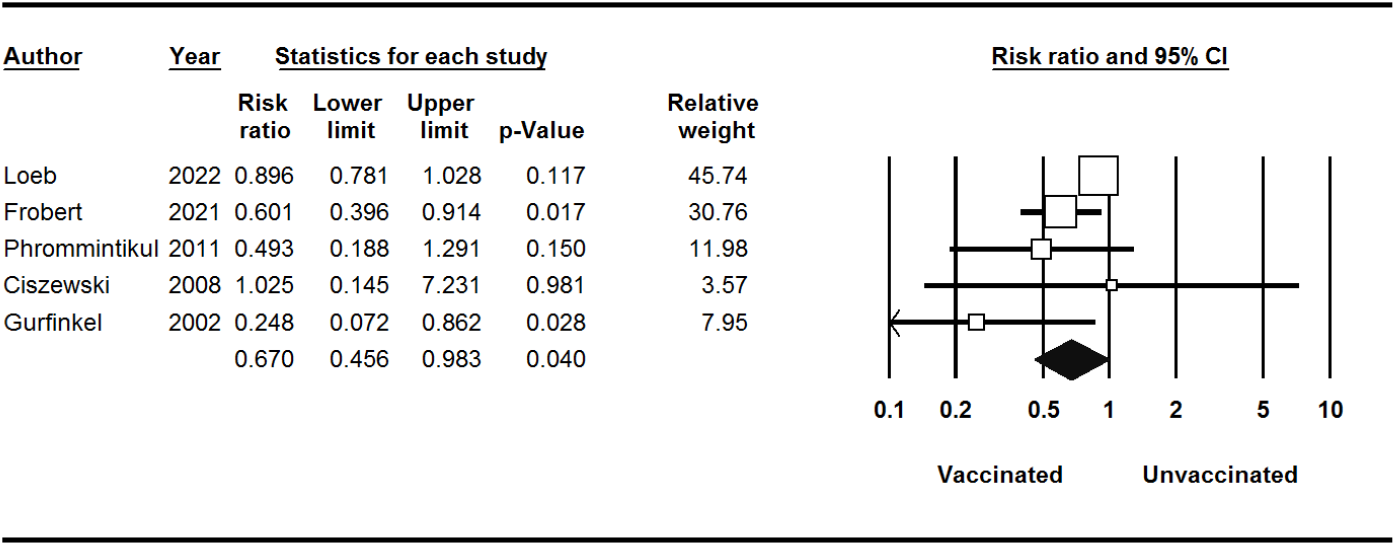
Cardiovascular Death Events for Influenza Vaccine Vs Control.

## Discussion

Revealing a compelling insight into the potential benefits of influenza vaccination, our comprehensive meta-analysis, based on the latest randomized controlled trial (RCT) data, demonstrates a significant interaction between influenza vaccination and the reduction of major cardiovascular events. Notably, patients who received the influenza vaccine experienced a remarkable risk reduction of over 20% in cardiovascular death. These findings underscore the potential impact of influenza vaccination in safeguarding against adverse cardiovascular outcomes among vulnerable patient populations.

### Comparisons with other studies

Our results with the Behrouzi et al. meta-analysis published in 2022 demonstrated a decreased risk of CVDs in patients receiving influenza vaccination [11].

However, our study has some strengths and is different in some ways. We also run stratified analyses based on MI, Stroke, and cardiovascular death. To minimize the influence of confounding factors, only studies that focused on a target population with pre-existing cardiovascular diseases (CVDs) were included in this analysis. Studies that involved sample populations with specific comorbidities were excluded from the analysis to ensure a more focused examination of the relationship between influenza vaccination and CVD outcomes. This approach aimed to isolate and better understand the direct impact of influenza vaccination on individuals with established cardiovascular conditions. Against this, we included one recent study that was not previously included. Our calculated RR for major cardiovascular events (0.70; 95%CI, 0.55-0.91) was a little higher than Behrouzi et al. (0.66; 95%CI, 0.53-0.83) but in line with its direction and statistical significance.

### Implications of the Findings

The observed interaction between influenza vaccination and reduced cardiovascular mortality among patients with recent CVDs is both clinically and epidemiologically significant. Cardiovascular diseases, including heart attacks and strokes, remain the leading causes of mortality worldwide. The potential of influenza vaccination to yield substantial reductions in cardiovascular mortality in this vulnerable patient group warrants serious attention from healthcare providers, policymakers, and researchers.

### Clinical Relevance and Public Health Impact

By targeting patients with recent CVDs for influenza vaccination, healthcare providers have a potential opportunity to mitigate the risk of cardiovascular death in a cost-effective and widely available manner. Influenza vaccination programs could be tailored to prioritize this high-risk group, thus potentially reducing the overall burden on healthcare systems and improving patient outcomes.

### Conclusion

The evidence from the most recent meta-analysis of RCT data is compelling, revealing a significant interaction between influenza vaccination and the reduction of major cardiovascular events among patients with recent CVDs. This finding underscores the potential benefits of targeting this high-risk group for vaccination. Further research is warranted to elucidate the precise mechanisms driving this association and to explore the long-term impact of influenza vaccination on cardiovascular outcomes. In the meantime, healthcare providers and policymakers should take heed of these findings and consider prioritizing influenza vaccination for patients with recent CVDs as a feasible and potentially life-saving preventive measure.

## Data Availability

All data produced in the present work are contained in the manuscript

## Notes

### Competing Interest Statement

The authors have declared no competing interest.

### Funding Statement

This study did not receive any funding

## References

1. Backer, J., et al., The impact of influenza vaccination on infection, hospitalisation and mortality in the Netherlands between 2003 and 2015. Epidemics, 2019. 26: p. 77–85.

2. Mameli, C., et al., Influenza vaccination: Effectiveness, Indications and Limits in the Pediatric Population. Frontiers in pediatrics, 2019. 7: p. 317.

3. Vamos, E.P., et al., Effectiveness of the influenza vaccine in preventing admission to hospital and death in people with type 2 diabetes. CMAJ, 2016. 188(14): p. E342–E351.

4. Joseph, P., et al., Reducing the global burden of cardiovascular disease, part 1: the epidemiology and risk factors. Circulation research, 2017. 121(6): p. 677–694.

5. Phrommintikul, A., et al., Influenza vaccination reduces cardiovascular events in patients with acute coronary syndrome. European heart journal, 2011. 32(14): p. 1730–1735.

6. Hsu, S.-Y., et al., A matched influenza vaccine strain was effective in reducing the risk of acute myocardial infarction in elderly persons: a population-based study. Medicine, 2016. 95(10).

7. DerSimonian, R. and N. Laird, Meta-analysis in clinical trials. Controlled clinical trials, 1986. 7(3): p. 177–188.

8. Loeb, M., et al., Randomized controlled trial of influenza vaccine in patients with heart failure to reduce adverse vascular events (IVVE): Rationale and design. Am Heart J, 2019. 212: p. 36–44.

9. Phrommintikul, A., et al., Influenza vaccination reduces cardiovascular events in patients with acute coronary syndrome. Eur Heart J, 2011. 32(14): p. 1730–5.

10. Ciszewski, A., et al., The protective effect of influenza vaccination on the clinical course of coronary disease in patients with acute coronary syndromes treated by primary PCI - A report from FLUCAD study. Postepy w Kardiologii Interwencyjnej, 2010. 6(1): p. 6–11.

11. Ciszewski, A., et al., Influenza vaccination in secondary prevention from coronary ischaemic events in coronary artery disease: FLUCAD study. Eur Heart J, 2008. 29(11): p. 1350–8.

12. Gurfinkel, E.P., et al., Flu vaccination in acute coronary syndromes and planned percutaneous coronary interventions (FLUVACS) Study. Eur Heart J, 2004. 25(1): p. 25–31.

13. Gurfinkel, E.P., et al., Influenza vaccine pilot study in acute coronary syndromes and planned percutaneous coronary interventions: the FLU Vaccination Acute Coronary Syndromes (FLUVACS) Study. Circulation, 2002. 105(18): p. 2143–7.

14. Christiansen, C.F., et al., Influenza vaccination and 1-year risk of myocardial infarction, stroke, heart failure, pneumonia, and mortality among intensive care unit survivors aged 65 years or older: a nationwide population-based cohort study. Intensive Care Med, 2019. 45(7): p. 957–967.

15. Wu, H.H., et al., Influenza vaccination and secondary prevention of cardiovascular disease among Taiwanese elders—A propensity score-matched follow-up study. PLoS ONE, 2019. 14(7).

16. Lavallee, P.C., et al., Influenza vaccination and cardiovascular risk in patients with recent TIA and stroke. Neurology, 2014. 82(21): p. 1905–13.

17. Johnstone, J., et al., Influenza vaccination and major adverse vascular events in high-risk patients. Circulation, 2012. 126(3): p. 278–86.

18. Hung, I.F., et al., Prevention of acute myocardial infarction and stroke among elderly persons by dual pneumococcal and influenza vaccination: a prospective cohort study. Clin Infect Dis, 2010. 51(9): p. 1007–16.

19. Gurfinkel, E.P. and R.L. de la Fuente, Two-year follow-up of the FLU Vaccination Acute Coronary Syndromes (FLUVACS) Registry. Tex Heart Inst J, 2004. 31(1): p. 28–32.

20. Macintyre, C.R., et al., Ischaemic heart disease, influenza and influenza vaccination: a prospective case control study. Heart, 2013. 99(24): p. 1843–8.

21. Heffelfinger, J.D., et al., Influenza vaccination and risk of incident myocardial infarction. Hum Vaccin, 2006. 2(4): p. 161–6.

22. Naghavi, M., et al., Association of influenza vaccination and reduced risk of recurrent myocardial infarction. Circulation, 2000. 102(25): p. 3039–45.

23. Meyers, D.G., et al., Influenza and pneumococcal vaccinations fail to prevent myocardial infarction. HeartDrug, 2004. 4(2): p. 96–100.

24. Chiang, M.H., et al., Association between influenza vaccination and reduced risks of major adverse cardiovascular events in elderly patients. Am Heart J, 2017. 193: p. 1–7.

25. Siriwardena, A.N., S.M. Gwini, and C.A. Coupland, Influenza vaccination, pneumococcal vaccination and risk of acute myocardial infarction: matched case-control study. CMAJ, 2010. 182(15): p. 1617–23.

## References

1. Treanor, J.J., Influenza vaccination. New England Journal of Medicine, 2016. 375(13): p. 1261–1268.

2. Azziz-Baumgartner, E., L. Grohskopf, and M. Patel, Realizing the potential of maternal influenza vaccination. Jama, 2021. 325(22): p. 2257–2259.

3. Wong, S.-S. and R.J. Webby, Traditional and new influenza vaccines. Clinical microbiology reviews, 2013. 26(3): p. 476–492.

4. Moher, D., et al., Preferred reporting items for systematic reviews and meta-analyses: the PRISMA statement. Annals of internal medicine, 2009. 151(4): p. 264–269.

5. Higgins, J.P., et al., The Cochrane Collaboration’s tool for assessing risk of bias in randomised trials. Bmj, 2011. 343.

6. Loeb, M., et al., Influenza vaccine to reduce adverse vascular events in patients with heart failure: a multinational randomised, double-blind, placebo-controlled trial. The Lancet Global Health, 2022. 10(12): p. e1835–e1844.

7. Fröbert, O., et al., Influenza vaccination after myocardial infarction: a randomized, double-blind, placebo-controlled, multicenter trial. Circulation, 2021. 144(18): p. 1476–1484.

8. Phrommintikul, A., et al., Influenza vaccination reduces cardiovascular events in patients with acute coronary syndrome. European heart journal, 2011. 32(14): p. 1730–1735.

9. Ciszewski, A., et al., Influenza vaccination in secondary prevention from coronary ischaemic events in coronary artery disease: FLUCAD study. European heart journal, 2008. 29(11): p. 1350–1358.

10. Gurfinkel, E.P., et al., Influenza vaccine pilot study in acute coronary syndromes and planned percutaneous coronary interventions: the FLU Vaccination Acute Coronary Syndromes (FLUVACS) Study. Circulation, 2002. 105(18): p. 2143–2147.

11. Behrouzi, B., et al., Association of influenza vaccination with cardiovascular risk: a meta-analysis. JAMA Network Open, 2022. 5(4): p. e228873–e228873.

