## Supplementary material for "Influenza Vaccination and Major Cardiovascular Risk: A Systematic Review and Meta-Analysis of Clinical Trials Studies": Table S1

Table S1. Search strategy for PubMed/Medline.

| **Search** | **Query** |
| --- | --- |
| #1 | (Myocardial Infarction[MeSH Terms]) OR Myocardial Infarction[Title/Abstract] OR Myocardial infarction [Title/Abstract] OR Cardiovascular[Title/Abstract] OR Atherosclerosis[Title/Abstract] OR Atrial fibrillation[Title/Abstract] OR Stroke[Title/Abstract] |
| #2 | (Influenza Vaccines[MeSH Terms]) OR Influenza Vaccine[Title/Abstract] |
| #3 | #1 AND #2 |
